# EMS prehospital response to the COVID-19 pandemic in the US: A brief literature review

**DOI:** 10.1101/2022.03.10.22272236

**Authors:** Christian Angelo I. Ventura, Edward E. Denton, Jessica A. David, Brianna J. Schoenfelder, Lillian Mela, Rebecca P. Lumia, Rachel B. Rudi, Barnita Haldar

**Author notes:** **CORRESPONDING AUTHOR:** (personal); Twitter: @venturachr. Submitted as a member of the Favored Author program.

## Abstract

This study aimed to analyze prehospital Emergency Medical Services (EMS) response to the COVID-19 pandemic in the US through a brief systematic review of available literature in context with international prehospital counterparts. An exploration of the NCBI repository was performed using a search string of relevant keywords which returned n=5128 results; articles that met the inclusion criteria (n=77) were reviewed and analyzed in accordance with PRISMA and PROSPERO recommendations. Methodical quality was assessed using critical appraisal tools, and the Egger’s test was used for risk of bias reduction upon linear regression analysis of a funnel plot. Sources of heterogeneity as defined by P < 0.10 or I^2 > 50% were interrogated. Findings were considered within ten domains: structural/systemic; clinical outcomes; clinical assessment; treatment; special populations; dispatch/activation; education; mental health; perspectives/experiences; and transport. Findings suggest, EMS clinicians have likely made significant and unmeasured contributions to care during the pandemic via nontraditional roles, i.e., COVID-19 testing and vaccine deployment. EMS plays a critical role in counteracting the COVID-19 pandemic in addition to the worsening opioid epidemic, both of which disproportionately impact patients of color. As such, being uniquely influential on clinical outcomes, these providers may benefit from standardized education on care and access disparities such as racial identity. Access to distance learning continuing education opportunities may increase rates of provider recertification. Additionally, there is a high prevalence of vaccine hesitancy among surveyed nationally registered EMS providers. Continued rigorous investigation on the impact of COVID-19 on EMS systems and personnel is warranted to ensure informed preparation for future pandemic and infectious disease response.

## INTRODUCTION

Prehospital clinical care under the purview of EMS has been traditionally understudied in the United States, as illustrated by the scarcity of literature as compared to other allied health professions. Under the federal oversight of the National Highway Traffic Safety Administration (NHTSA), EMS models of care vary drastically, shaped largely by geographic assignment, local community partners, and additional presiding authorities. Common models include municipality-based, private nonprofit, and private for-profit services **(1)**.

Within these structural differences exist a subset variance of agencies’ scope of practice. In accordance with the National EMS Scope of Practice Model, the Basic Life Support (BLS) scope extends to Emergency Medical Responders (EMRs) and Emergency Medical Technicians (EMTs), while the Advanced Life Support (ALS) scope is characteristic of Advanced EMTs (AEMTs) and Paramedics (EMT-Ps) **(2)**. As of January 2022, COVID-19, the respiratory infection caused by novel SARS-CoV-2, is a leading cause of death in the US **(3)**. This global pandemic has established an unprecedented call for rigorous investigation of resource capacity and competency amid allied health care industries to ascertain structural deficits and strengths that have implications for future pandemic response. Further, preliminary mixed-method studies on EMS pandemic response in the context of the COVID-19 pandemic have identified deficits in the domains of resource availability, continued education, administrative protocols, and decontamination practices **(4)**.

Although evidently there is an apparent scarcity of academic literature centering US EMS systems in response to the COVID-19 pandemic, it may be useful to summatively review existing literature of satisfactory methodical quality to highlight research areas that warrant further investigatory attention, as no review of this scope and nature currently exists. In this study, we aimed to systematically review peer-reviewed literature indexed in PubMed pertaining to COVID-19 considerations, effects, and implications on US EMS systems and clinicians within the context of prehospital international counterparts by employing artificial intelligence and conventional PRISMA and PROSPERO recommendations.

## METHODS

### Search Strategy

On 07 March 2022 a PubMed National Center for Biotechnology Information (NCBI) repository search was conducted using the keywords “COVID,” “COVID-19,” “SARS-COV-2,” “CORONAVIRUS,” “EMERGENCY MEDICAL SERVICES,” “EMS”, “EMT”, “PREHOSPITAL,” “OUT OF HOSPITAL,” and “PARAMEDIC.” The returned English results were uploaded to Rayyan.ai for comprehensive abstract and full text review **(5)**.

### Data Extraction

Duplicate results were assessed for and removed. Editorials, commentaries, and non-peer reviewed manuscripts were excluded. Two investigators independently reviewed abstracts to identify articles eligible for full text review. The investigators then independently reviewed full text articles to identify studies that met the PICOS-guided inclusion criteria **(6)**. Studies were excluded if there were concerns regarding methodical quality or integrity of the data as per discretion of the two investigators and a third consultant. SIGN appraisal tools were used to exclude retrospective and cohort-based studies that did not meet an acceptable level of evidence for inclusion in the review **(7)**. Conflicts were resolved through discussion and by mediation from a third or fourth consultant when necessary. Study methods were consistent with PRISMA recommendations, and although PROSPERO registration was not sought, the work remained faithful to conventional review standards **(8,9)**.

### Analysis

Stata/BE software was used to assess aggregate prevalence of data. Heterogeneity was defined as P < 0.10 or I^2 > 50% which warranted a fixed effects approach **(10)**. Otherwise, a random effects approach was assumed. We then sought to investigate sources of heterogeneity. During full text review, studies were primarily taxonomized in one of the following ten domains: i. structural/systemic, ii. clinical outcomes, iii. clinical assessment, iv. treatment, v. special populations, vi. dispatch/activation, vii. education, viii. mental health, ix. perspectives/experiences, x. transport.

### Methodical Quality

Investigators determined risk of bias for prevalence studies based upon appropriateness of sampling, sampling methods, use of standard assessment methods, and setting characteristics. The criteria were adapted from either the Joanna Briggs Institute Critical Appraisal Checklist for Studies Reporting Prevalence Data or the Joanna Briggs Institute Critical Appraisal Tool for Quasi-Experimental Studies **(11,12)**. A 95% confidence interval was used to standardize prevalence and pooling of data, and the linear regression based Egger’s test was used to analyze the presence of any publication bias after a funnel plot was created and assessed **(13)**. P > 0.05 deemed no risk of publication bias. Microsoft Excel was used to calculate standard deviation and mean. Sensitivity analyses assessed for source of heterogeneity and stability of results. Because the work did not involve the use of human research subjects, it did not require approval or review by an institutional review board or bioethics committee.

### Diversity in the Research Team

The work represents a cross-institutional and multi-disciplinary collaboration amongst public health oriented researchers, EMS clinicians, and physician scientists.

## RESULTS

The NCBI repository returned n=5128 results, with n=77 utilized for analysis and inclusion in this study. Results were excluded if they did not satisfy the inclusion criteria. Figure 1 depicts an overview of the exclusion schema. AI identified n=111 duplicative results, n=4916 results were excluded due to irrelevance with respect to the area of investigation, and n=24 studies were excluded after investigators performed full text reviews and found studies to be of unsatisfactory evidence levels in accordance with SIGN appraisal guidelines. Table 1 depicts characteristics of studies selected for inclusion, with high confidence in stability and quality of the aggregate data.

**Figure 1.**
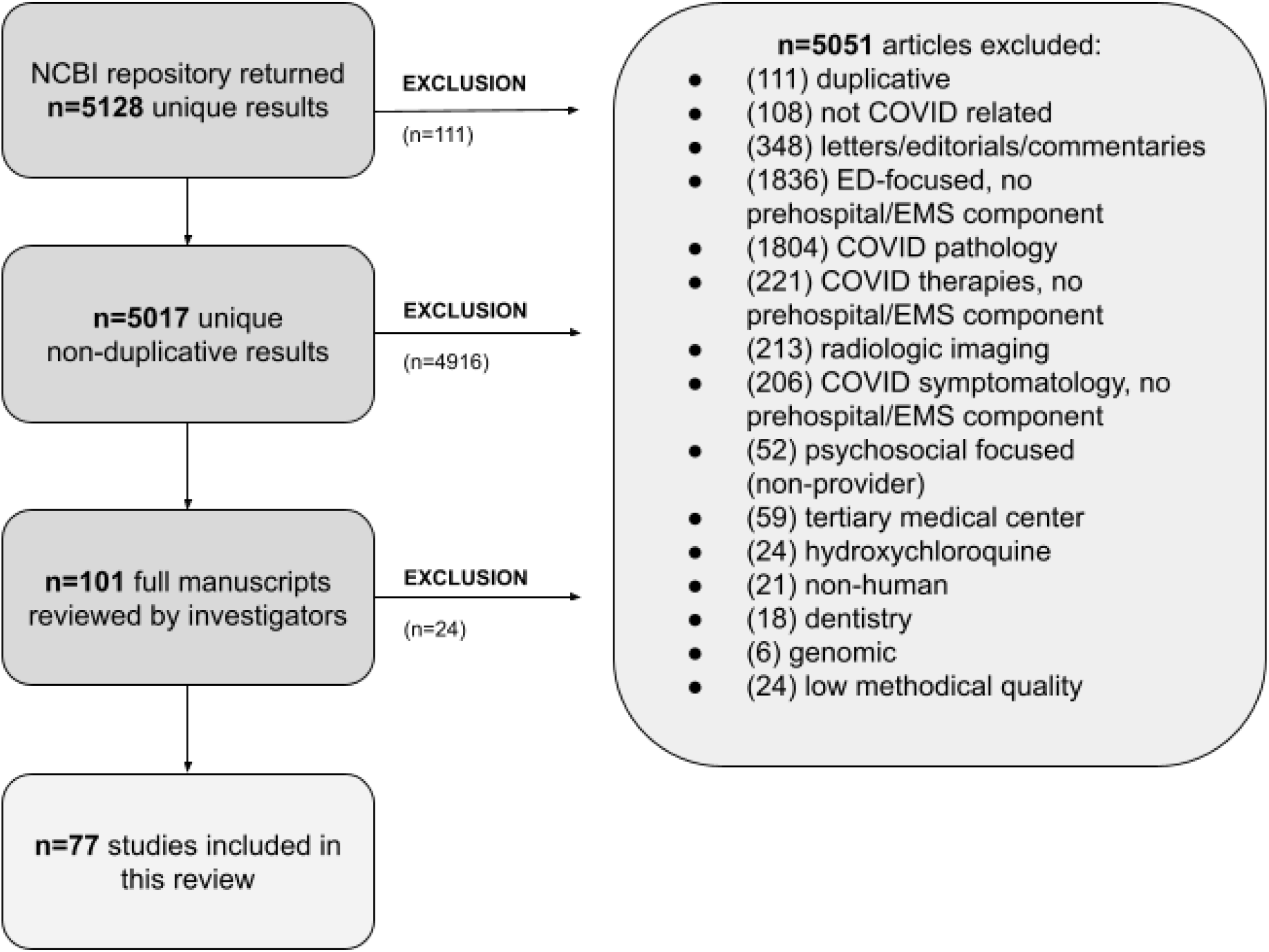
Schematic selection of relevant articles for inclusion in review.

**Table 1.**
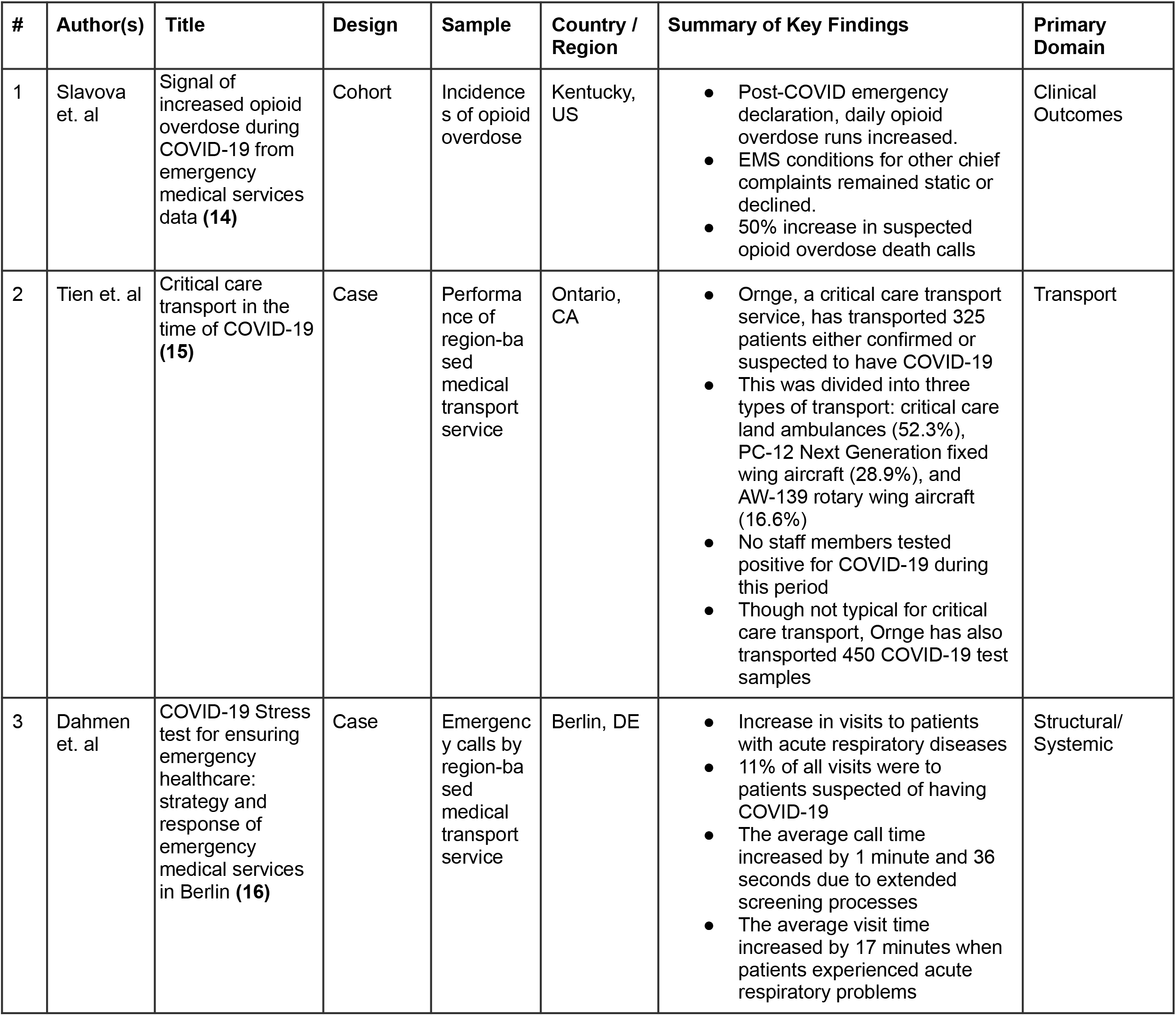
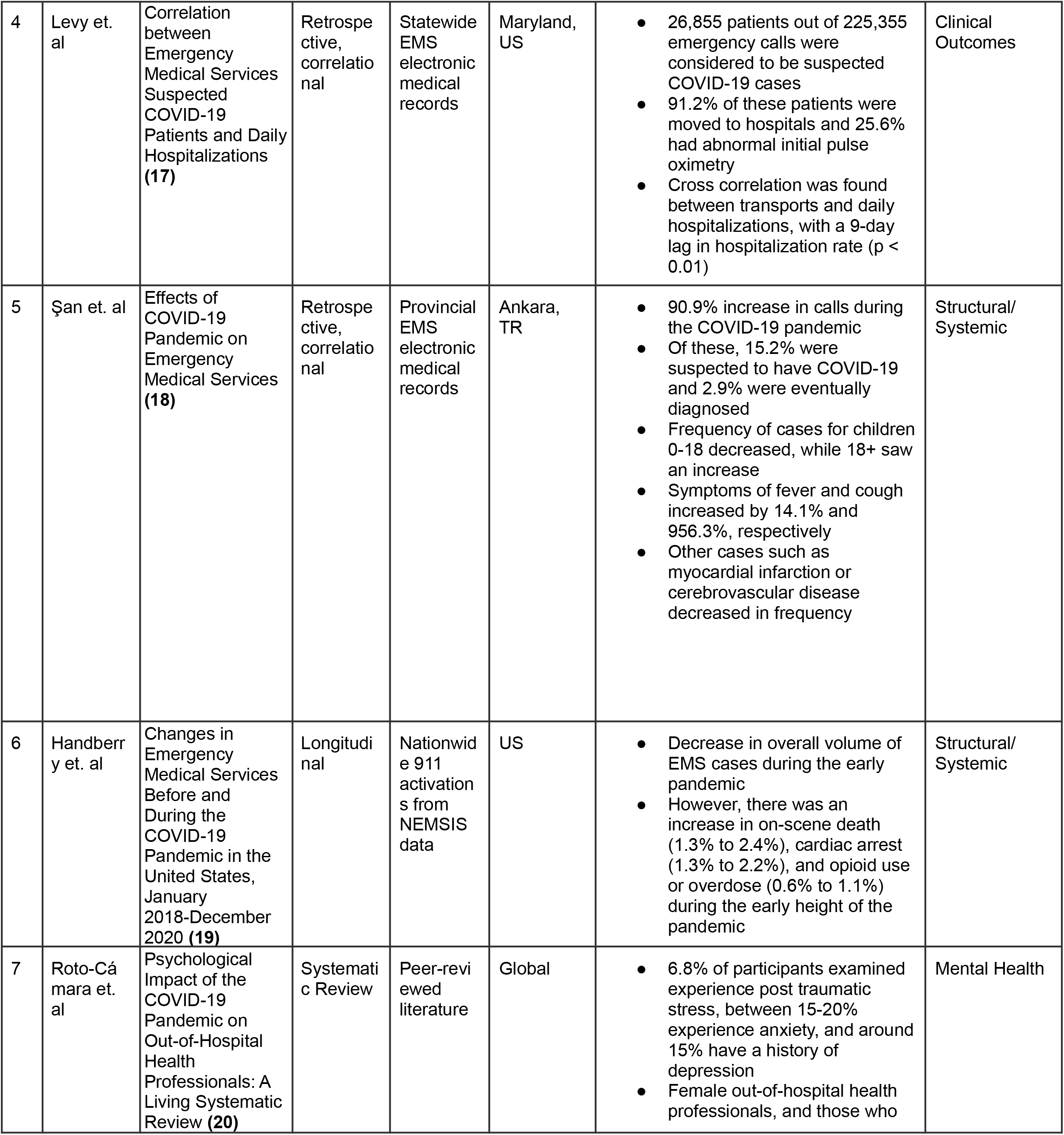
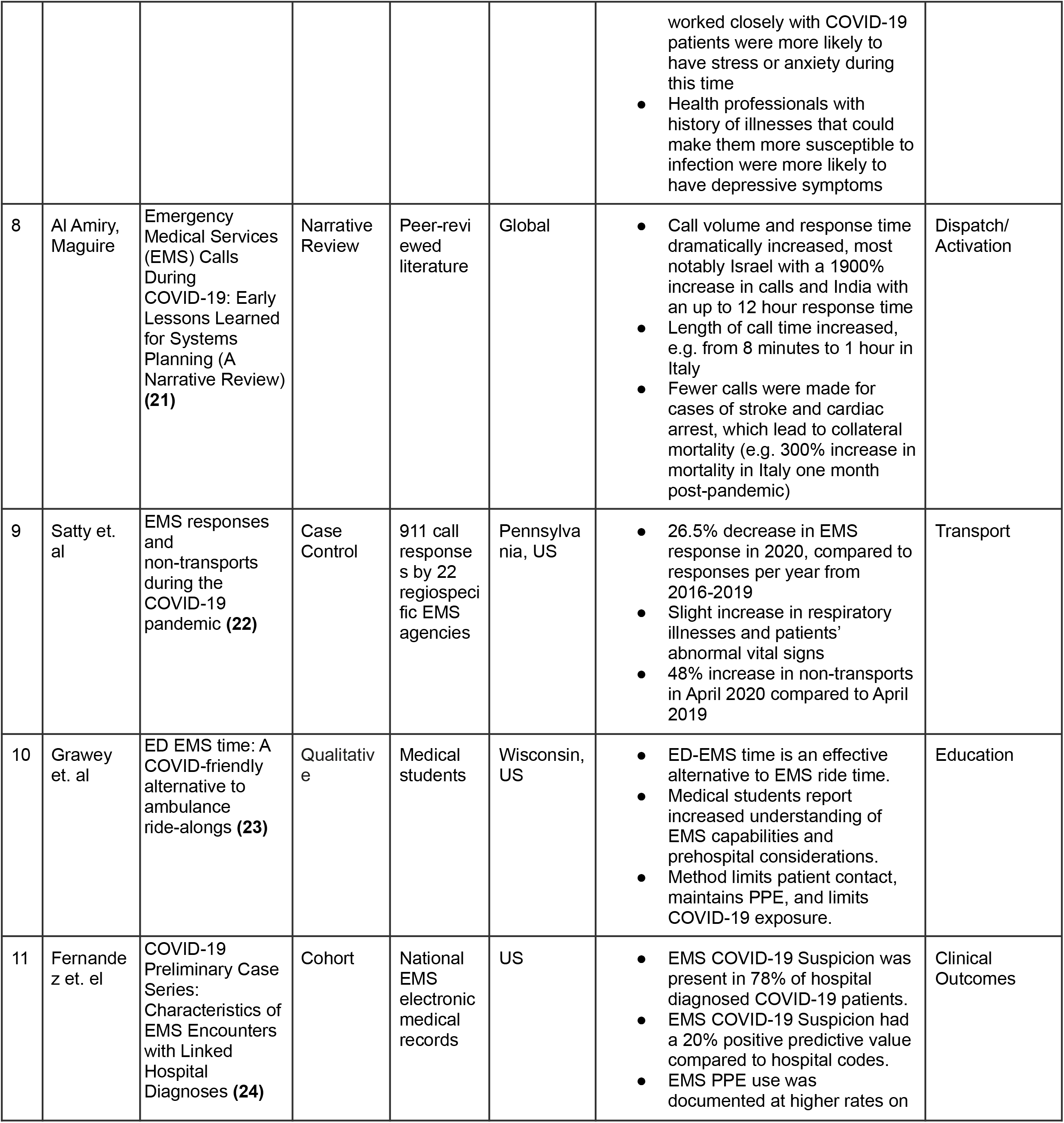
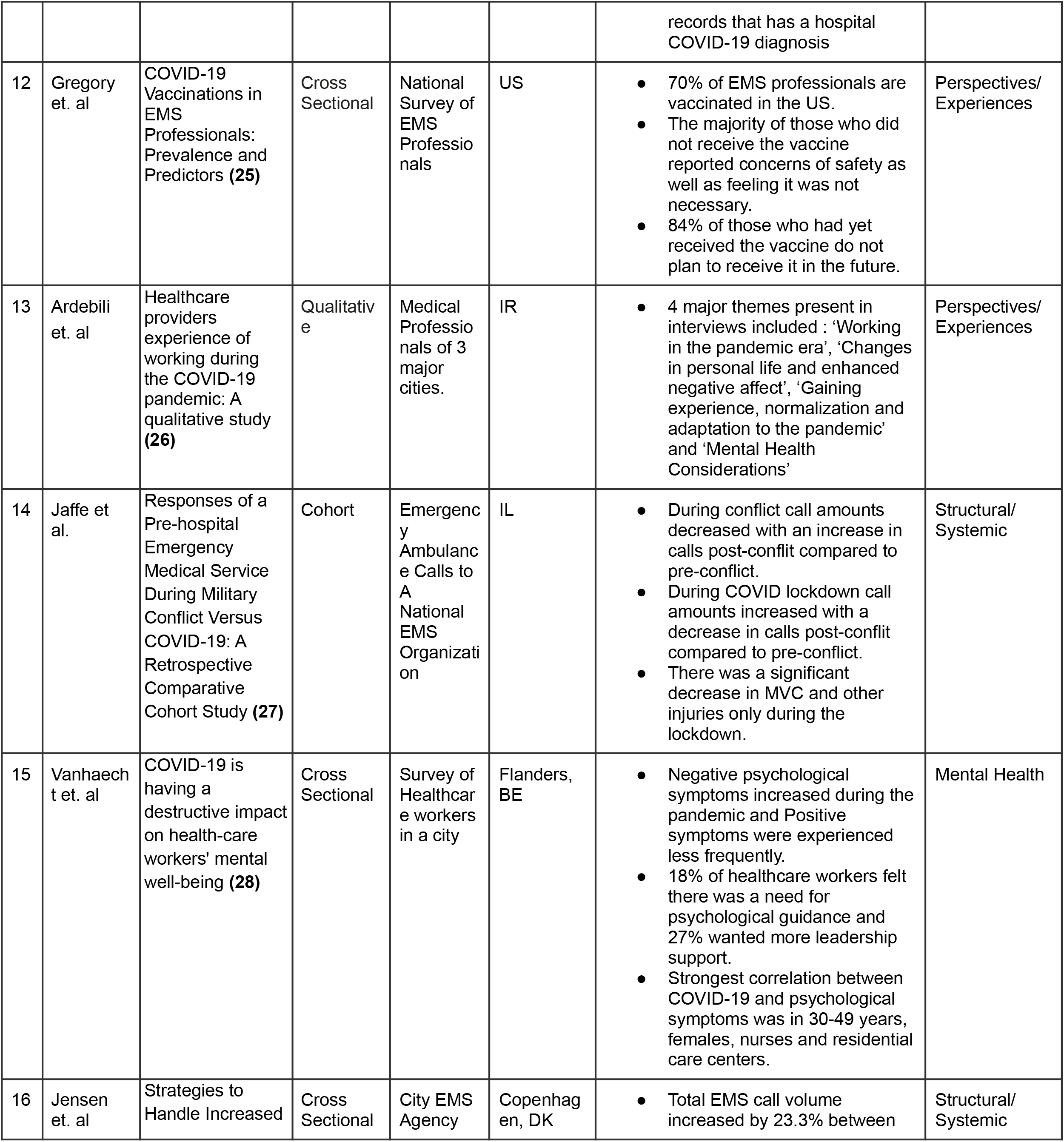
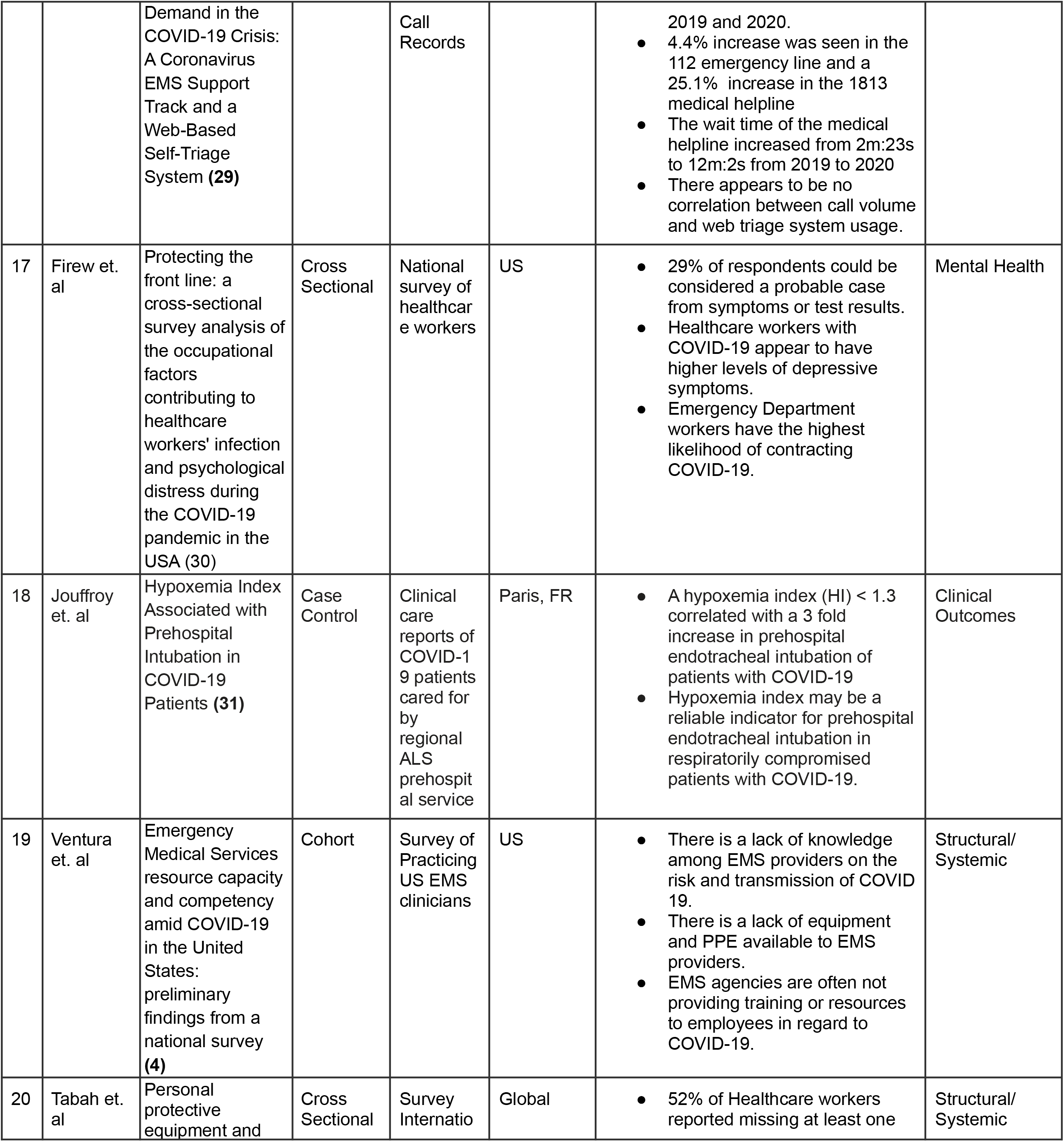
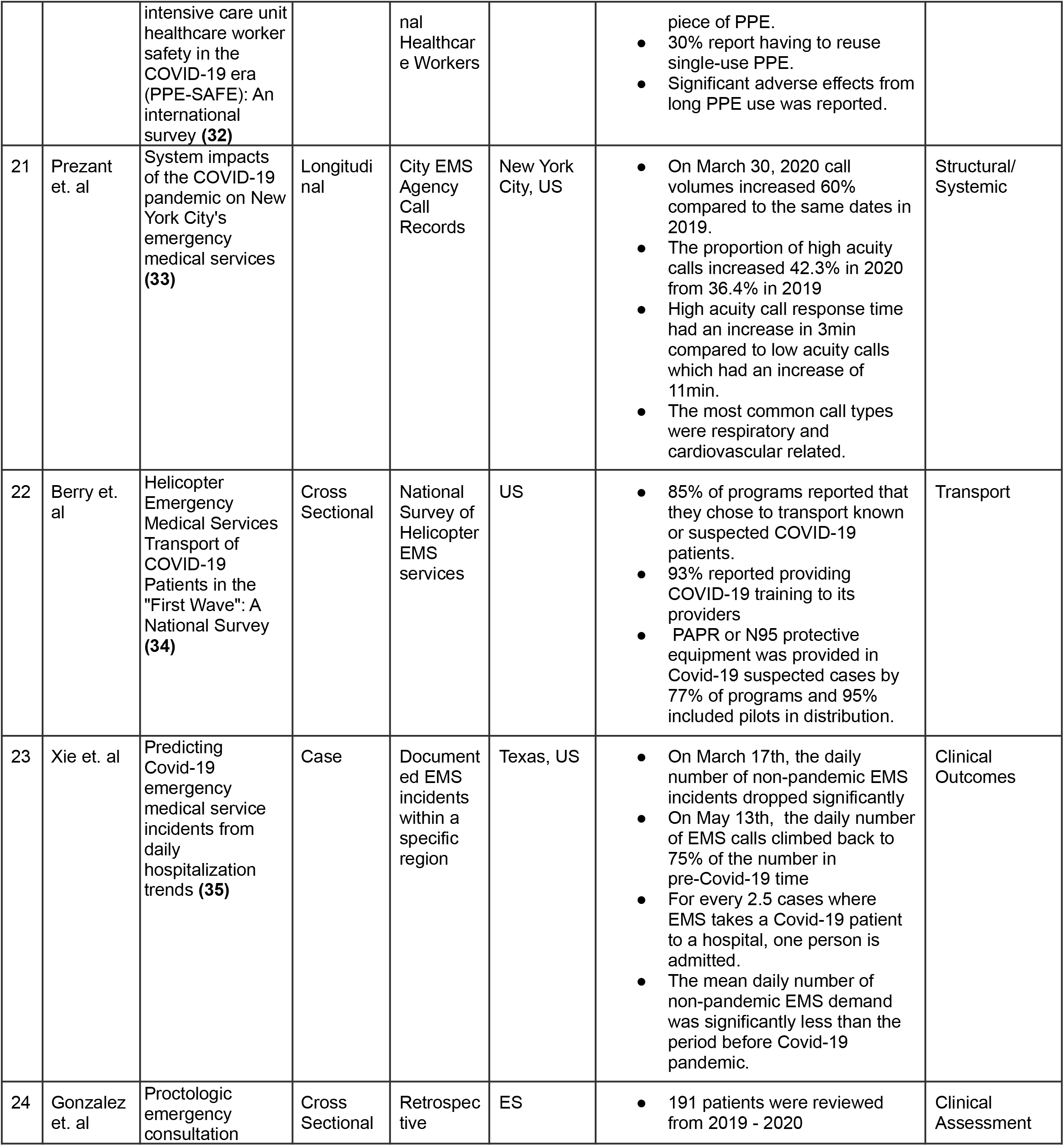
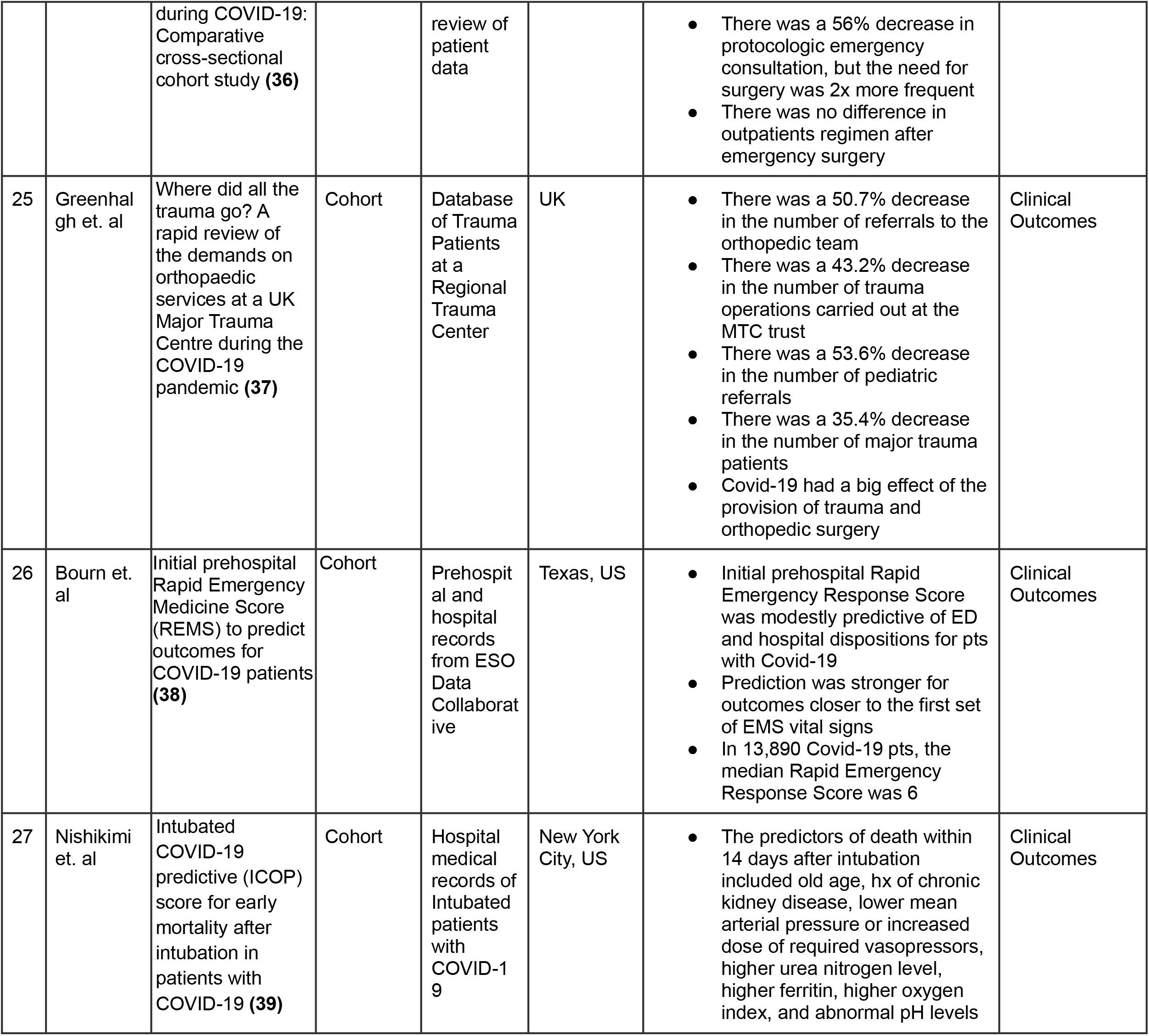
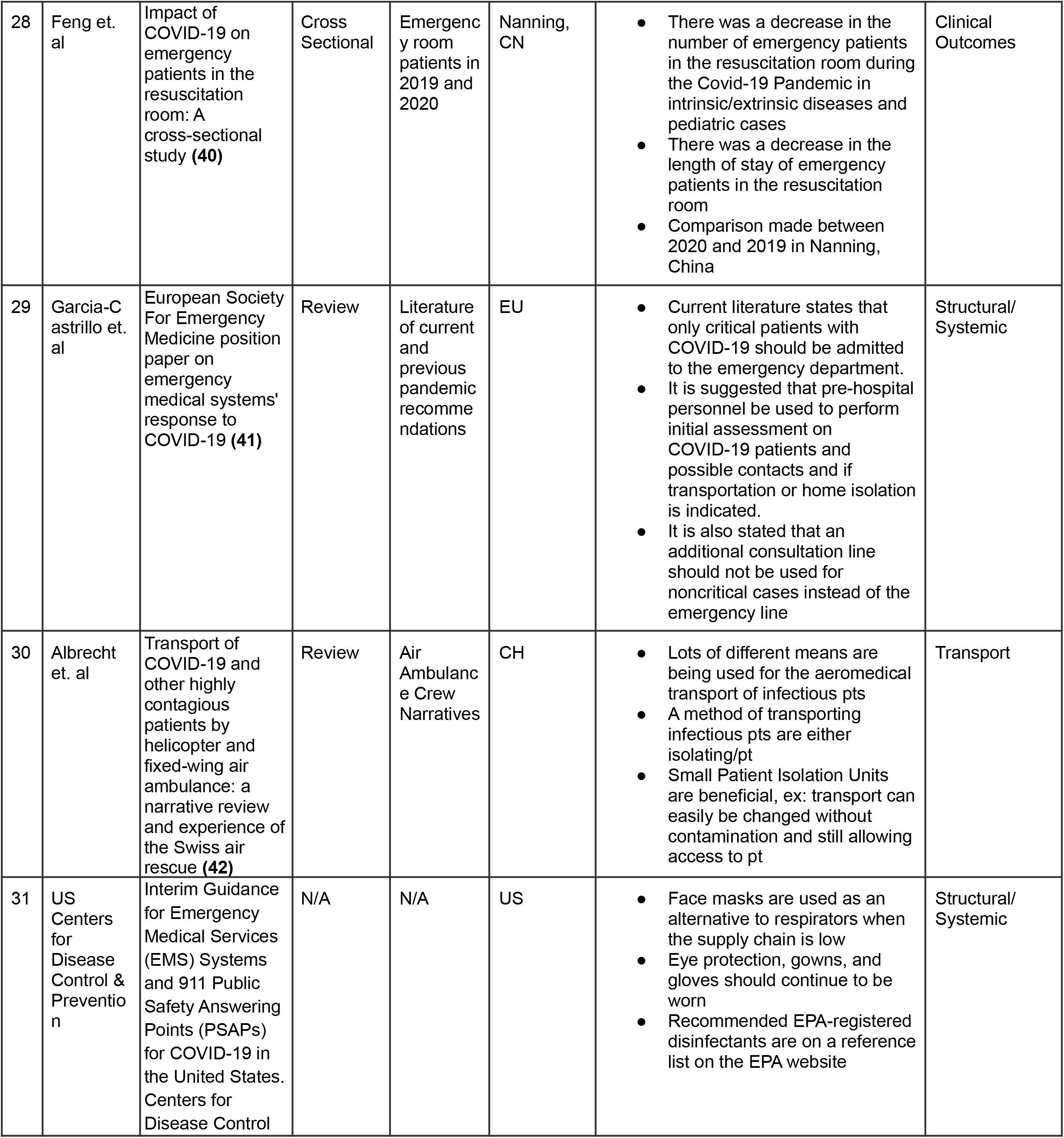
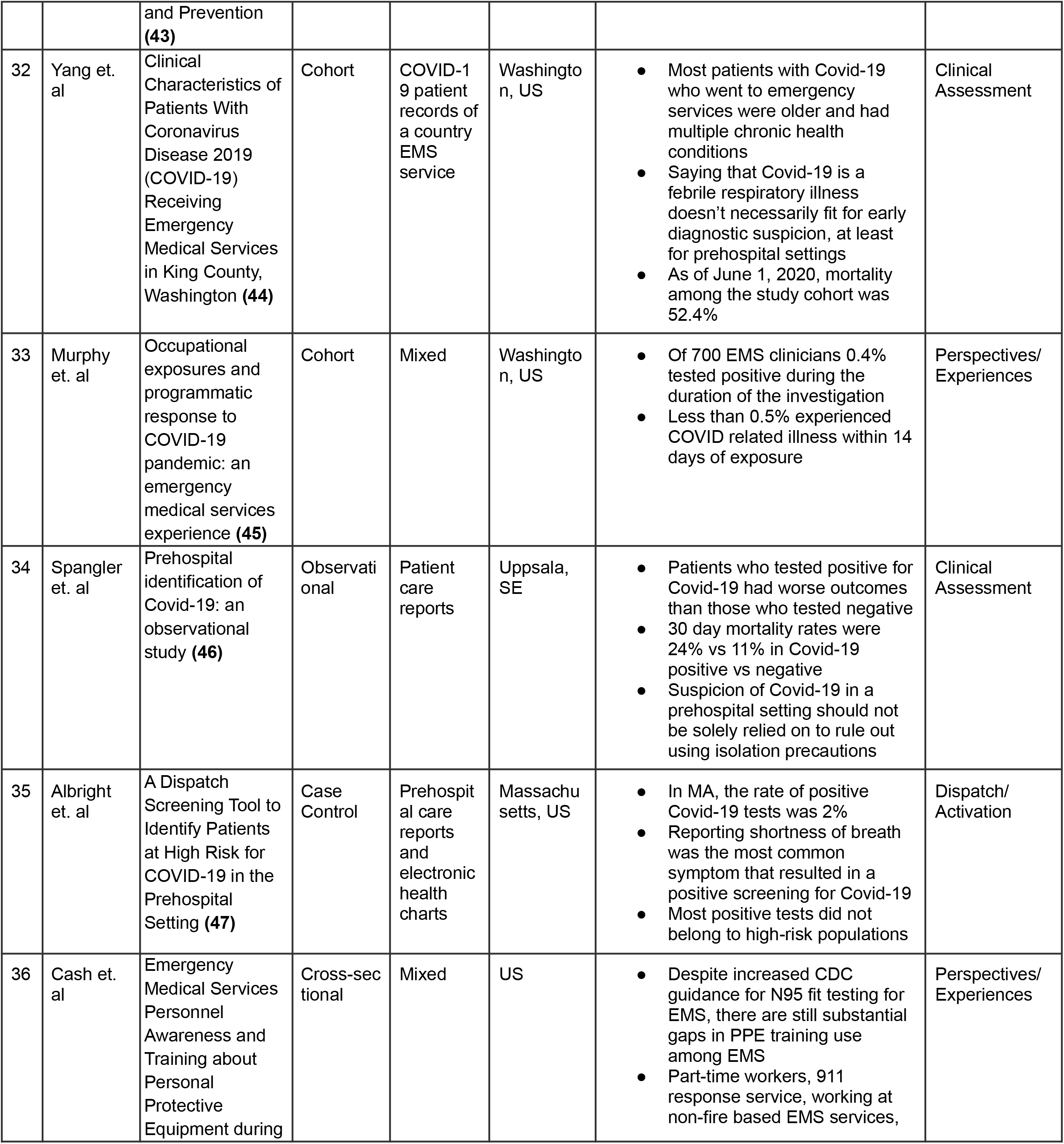
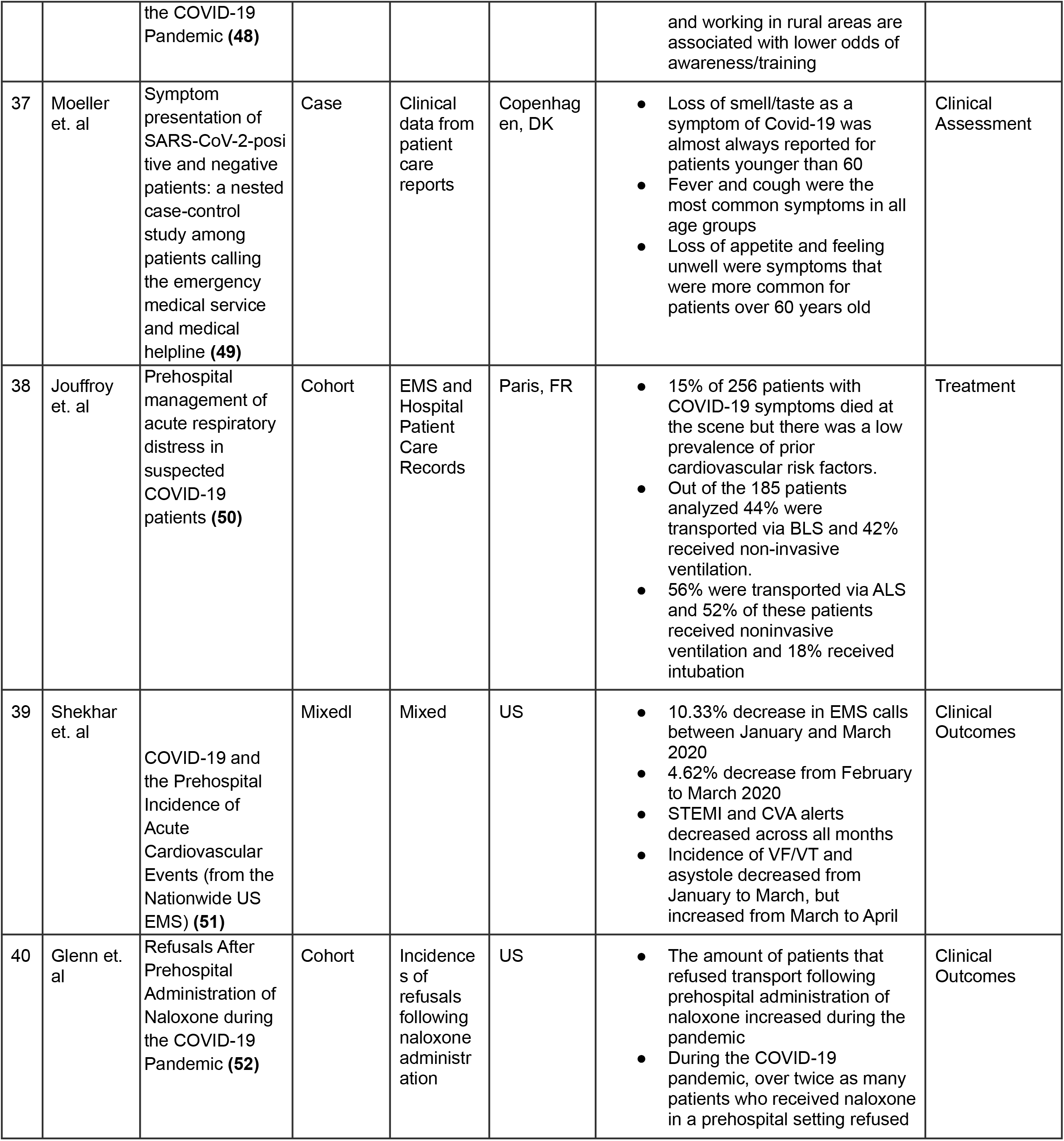
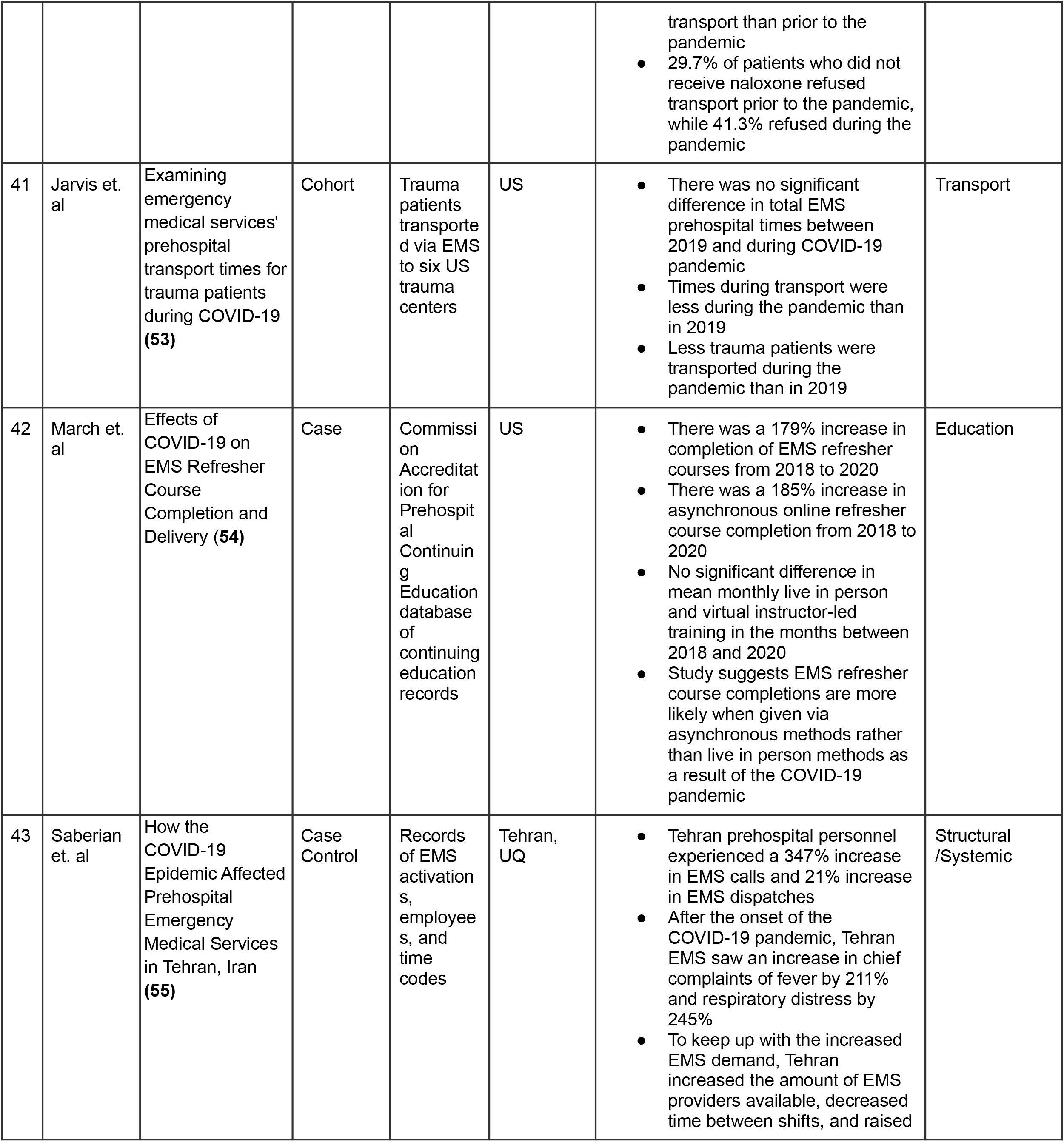
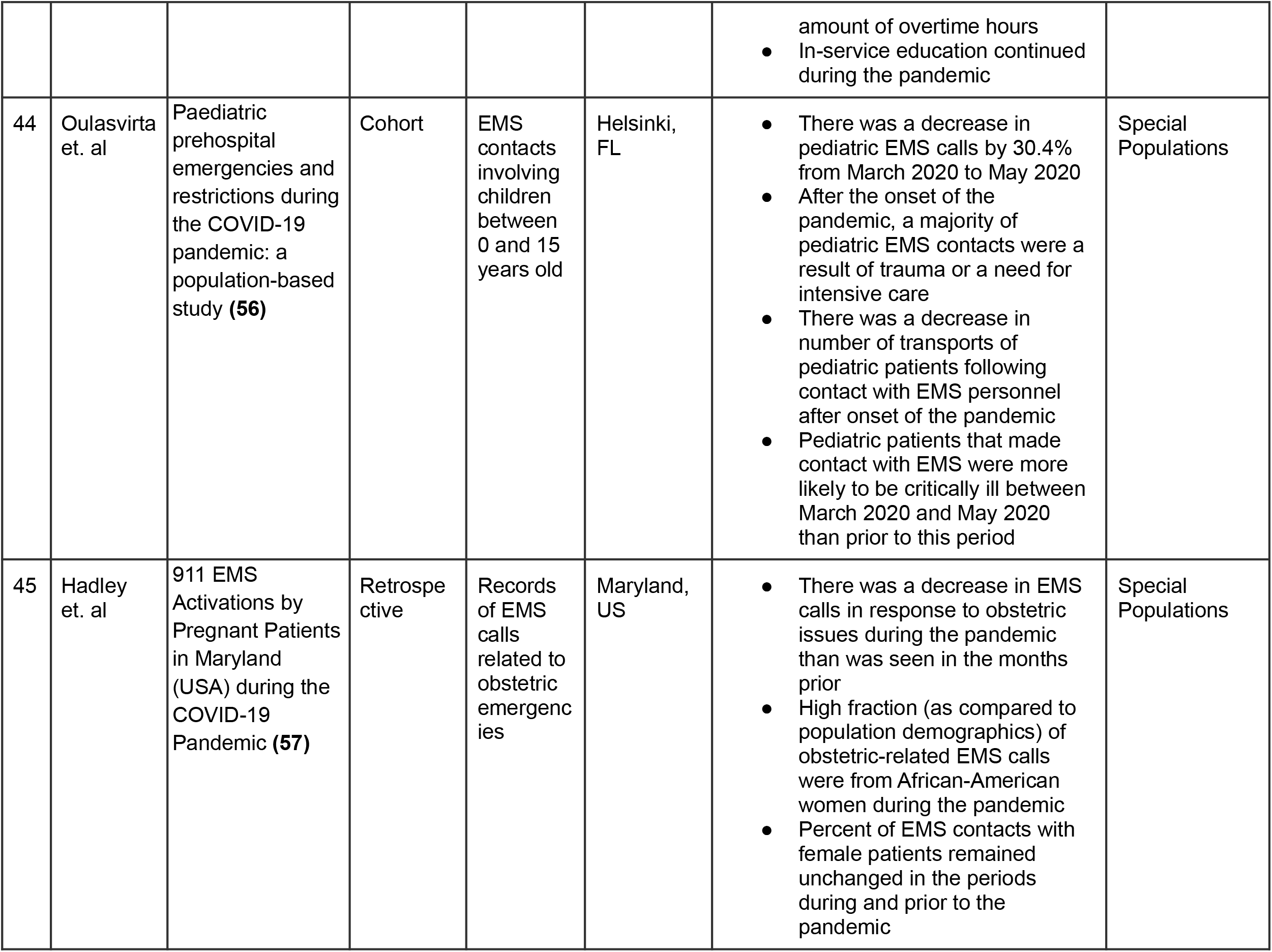
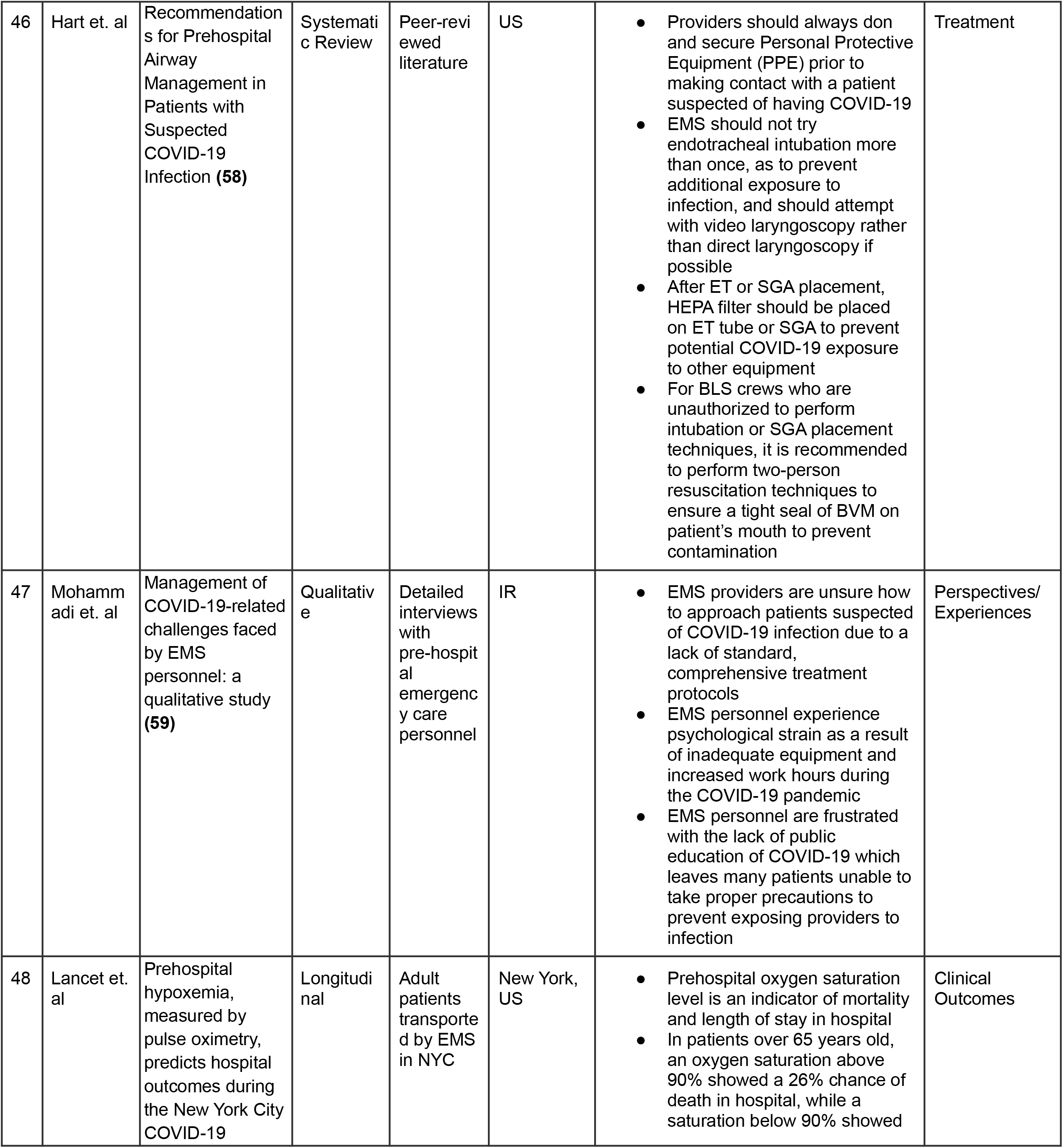
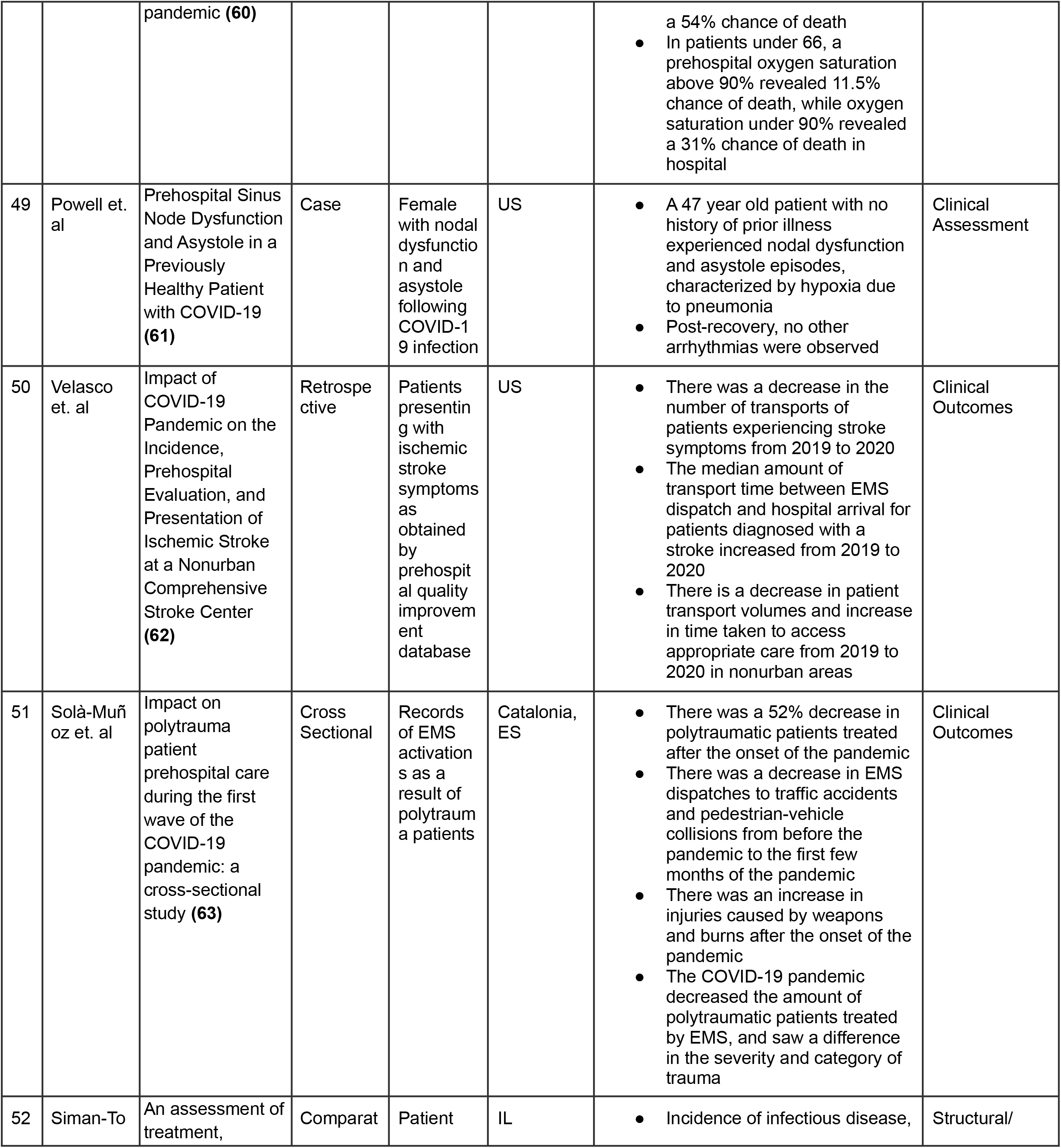
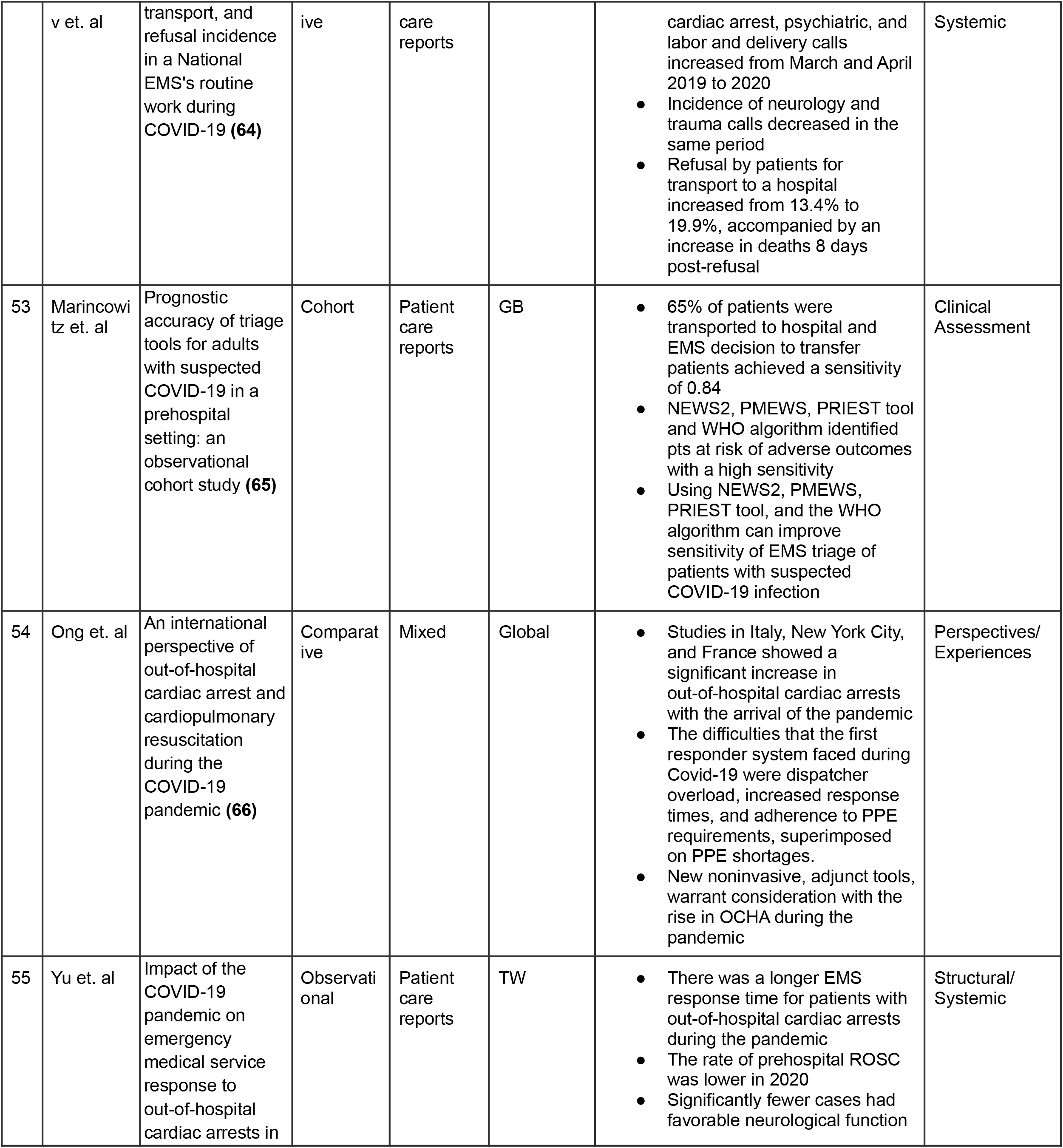
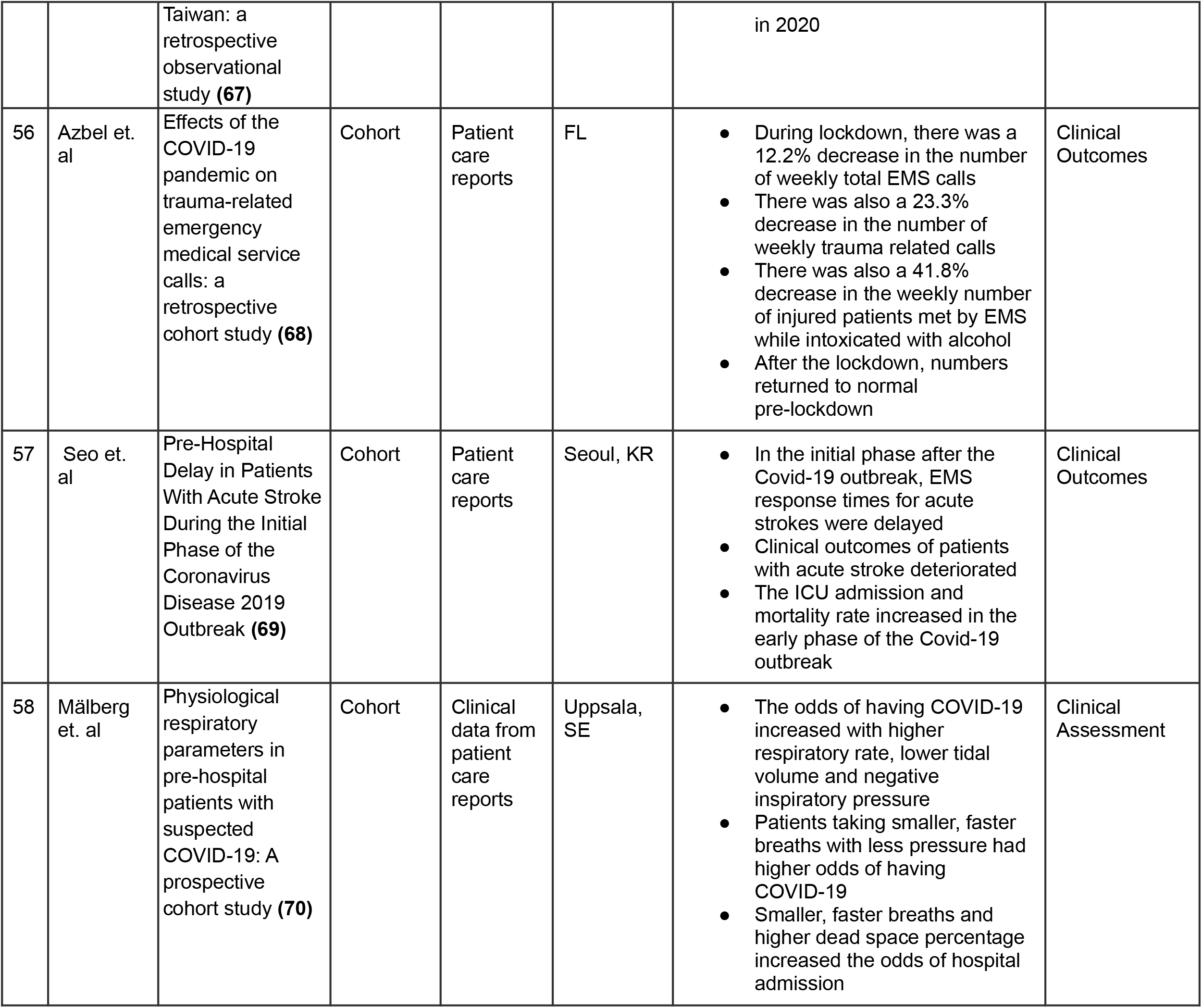
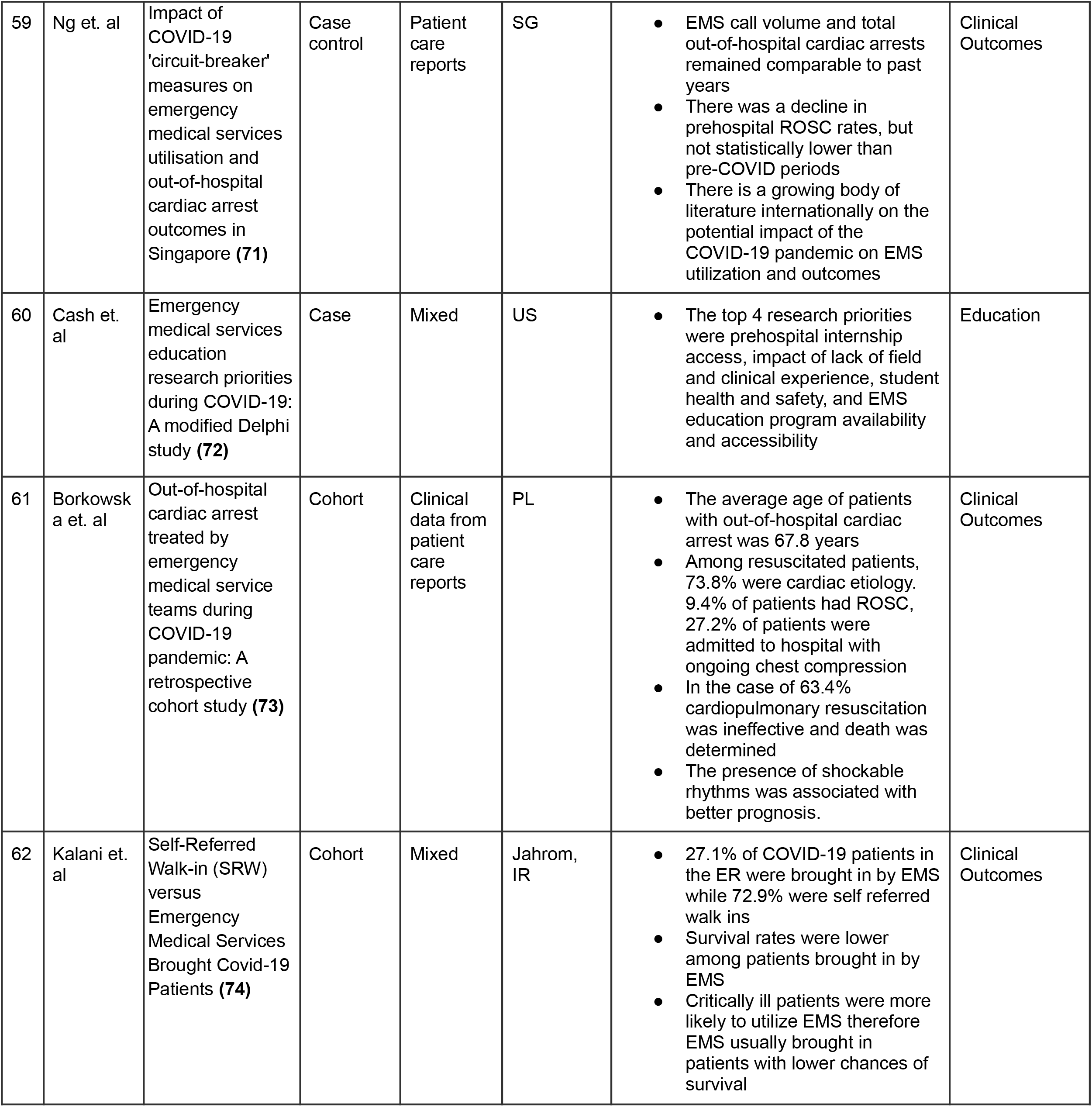
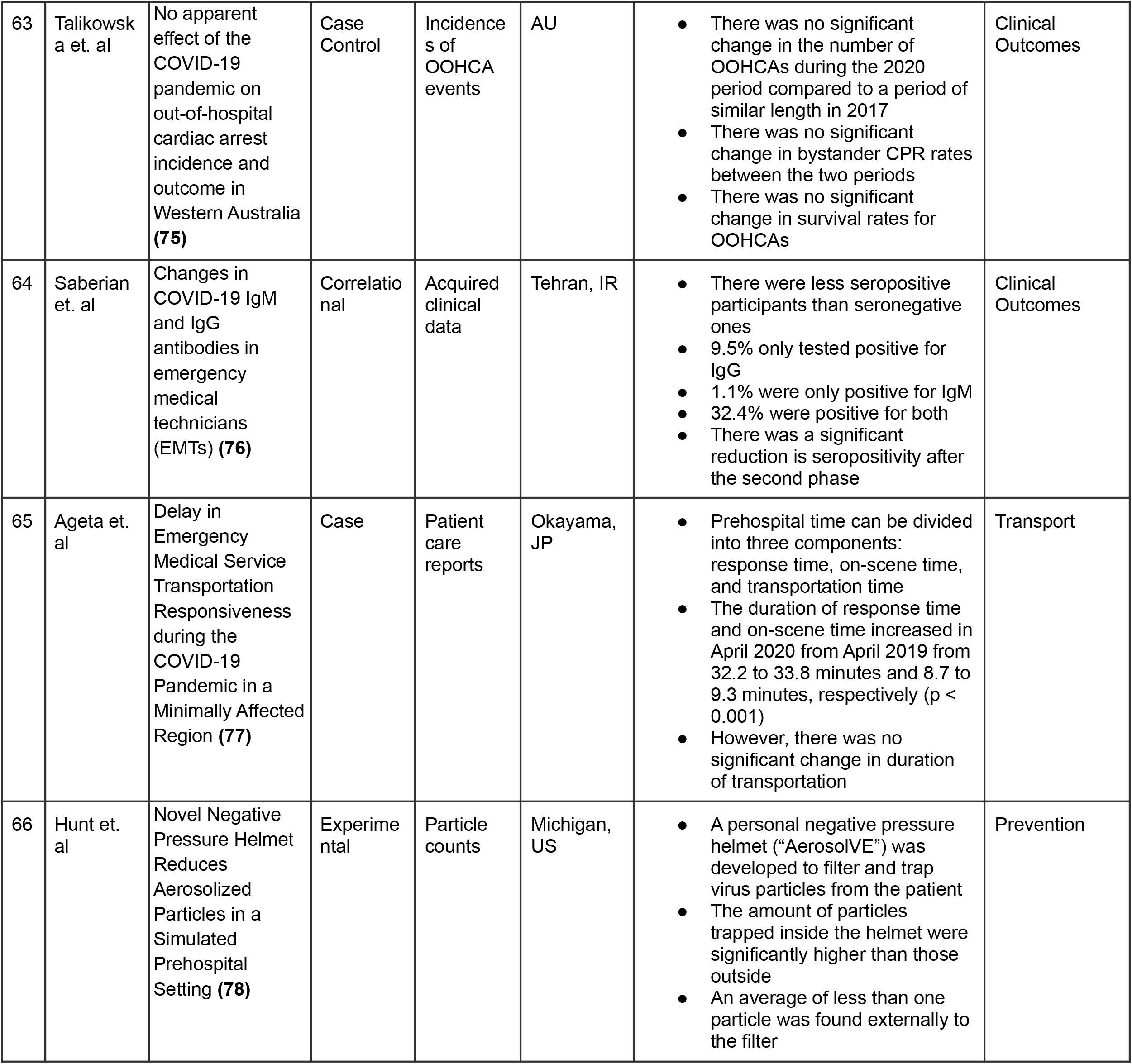
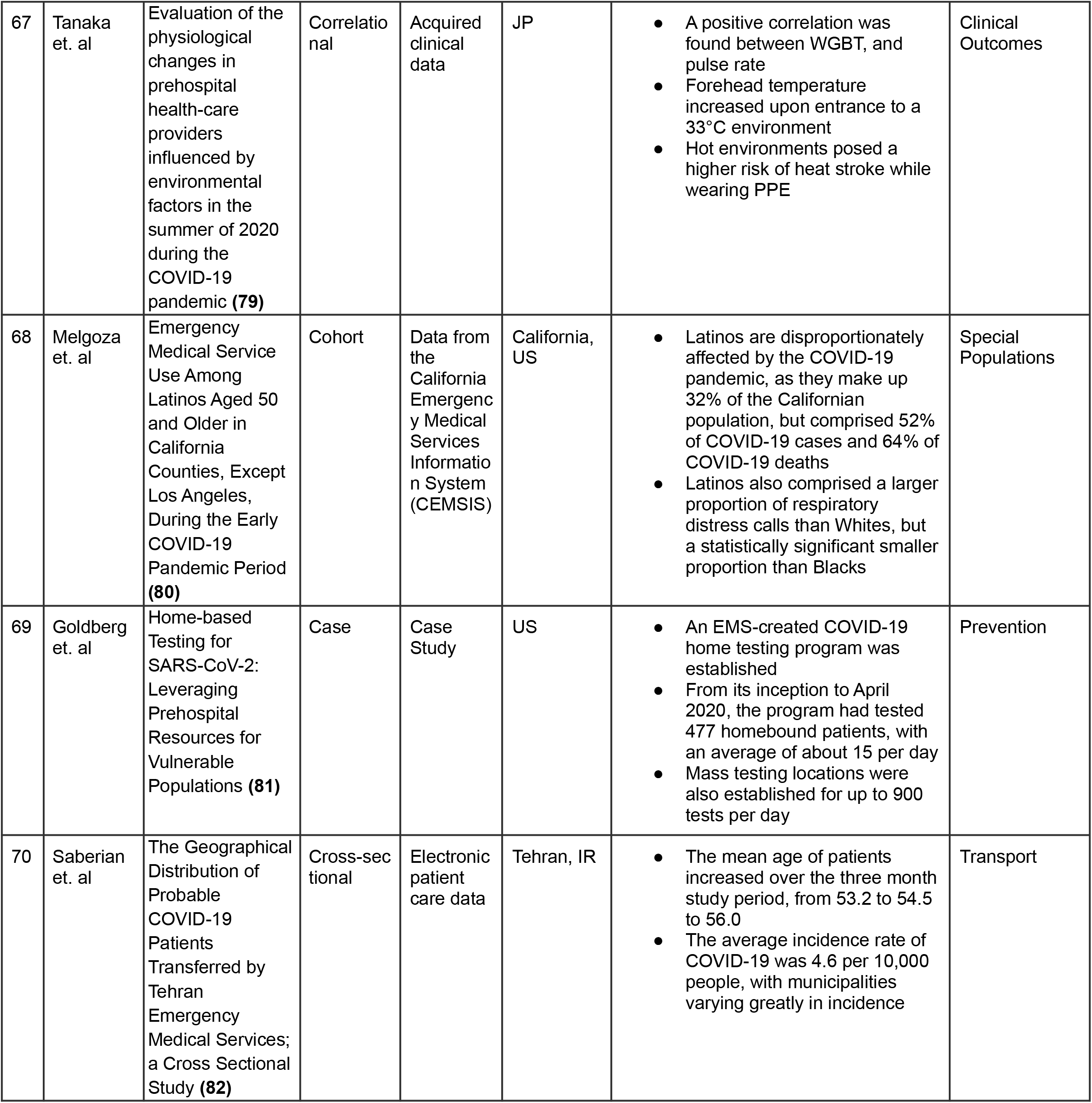
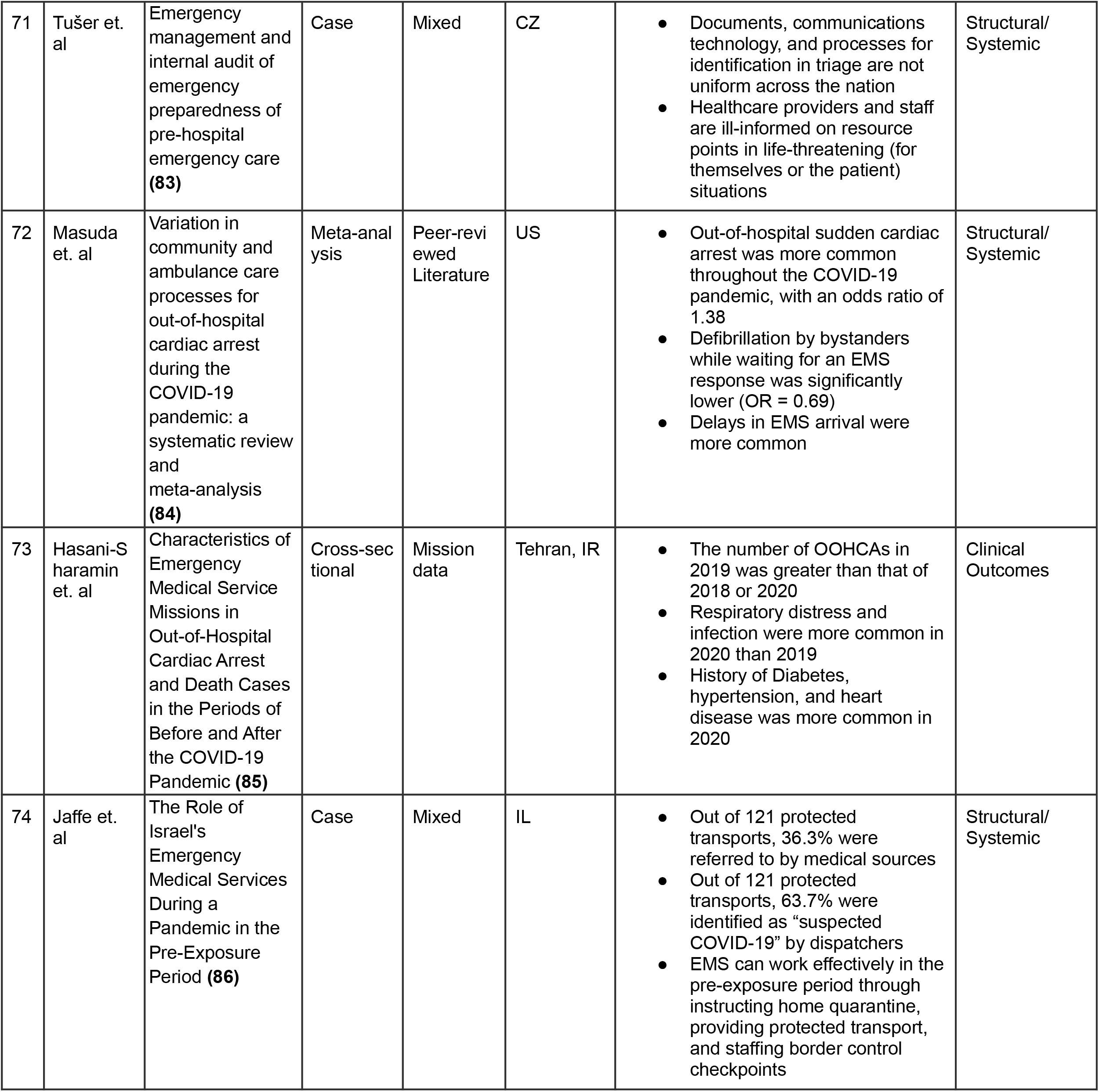
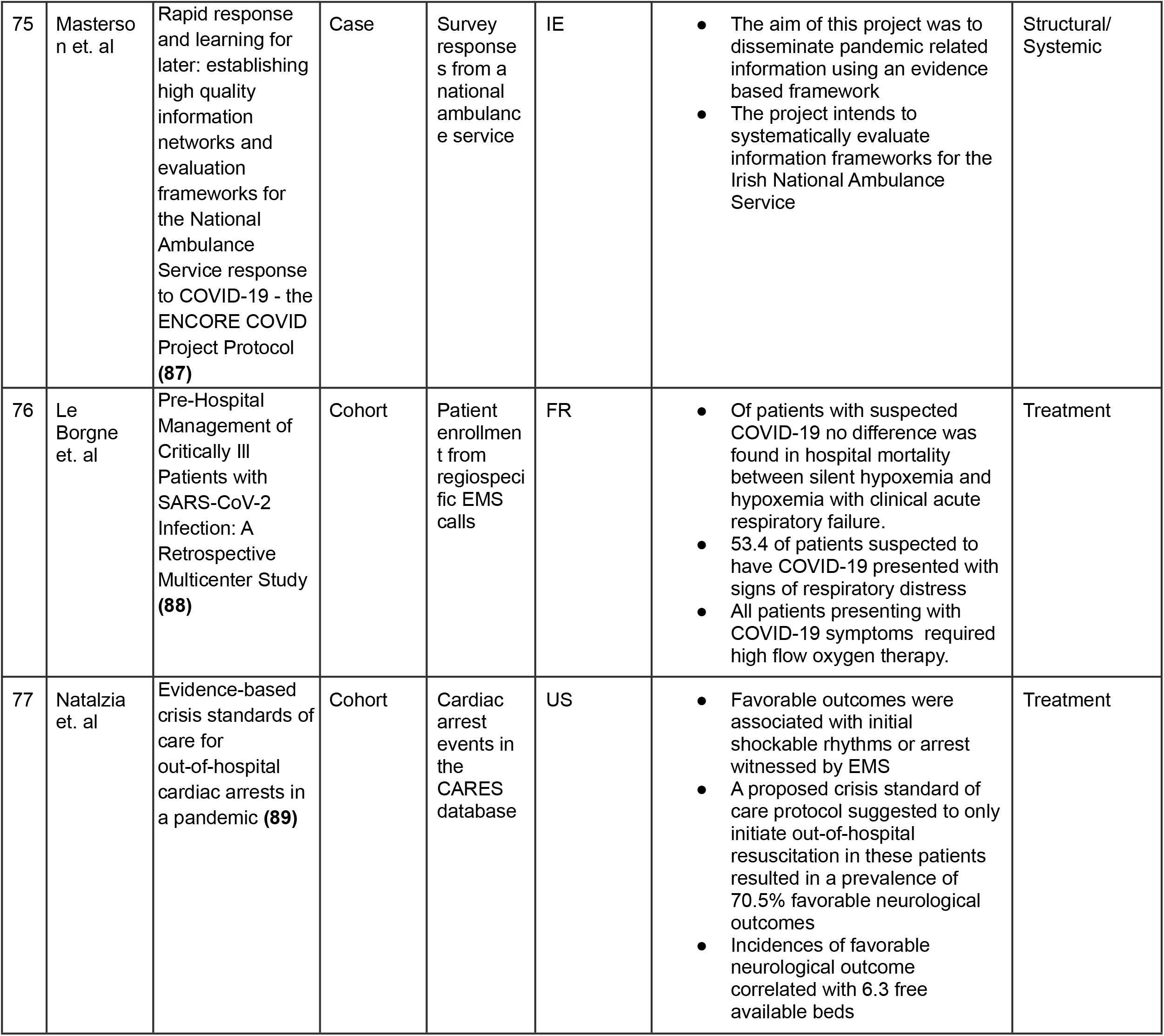
Characteristics of studies selected for inclusion

Of the included studies, 44% were US-based. Primary taxonomy schema revealed the following prevalence of data: n=16 structural/systemic, n=23 clinical outcomes, n=7 clinical assessment, n=4 treatment, n=3 special populations, n=2 dispatch/activation, n=6 education, n=3 mental health, n=6 perspectives/experiences, and n=7 transport (see Table 1).

## DISCUSSION

### Structural Considerations

Of the studies selected for inclusion in the “structural/systemic” domain, only 25% pertained to regions within the US. Only one study at the start of the pandemic (April 2020) rigorously assessed resource capacity and clinical competency at the national level, in response to the COVID-19 pandemic. Study participants were licensed practicing EMS clinicians in all 50 US states including the District of Columbia. Ventura et. al report regular dissatisfactory decontamination practices both prior to and during the pandemic, required prolonged use of N95 respirators past the recommended efficacy period, and overall inadequate education for infection control and clinical considerations for COVID-19 patients **(4)**. Notably, this captures a vulnerable representation of the US EMS system when there was minimal consensus on COVID-19 clinical symptomatology, transmissibility, appropriate precautionary measures, and discovery of vaccines. Globally, highly populated regions in the US, Germany, Turkey, Ireland, Denmark observed an increase in emergency medical calls of varying chief complaints, with many agency and regiospecific ambulance agencies reporting call volumes exceeding 60% compared to 2019 **(14,16,33)**. Based upon analysis of 911 de-identified patient care reports from the National EMS Information System (NEMSIS), between January 2018 to December 2020, Handberry et. al report an increase in frequency of on-scene death (1.3% to 2.4%), cardiac arrest (1.3% to 2.2%), and opioid related emergencies (0.6% to 1.1%) at the national level **(6)**. States such as Kentucky observed higher rates of opioid related 911 calls, with a 50% increase in suspected opioid overdose death calls, while calls for other chief complaints remained static or declined **(14)**. This is consistent with an apparent national trend of a worsening opioid epidemic, which we hypothesize has been only exacerbated by COVID-19. In February 2022, the American Medical Association issued a statement alleging an increase in drug related deaths in all US states and territories **(90)**. To that end, it is critical to acknowledge that recent studies suggest black and brown communities are likely to be disproportionately harmed by both the opioid epidemic and the COVID-19 pandemic, as rates of morbidity and mortality are generally higher in this clinical population **(91,92,93,94)**. Because EMS clinicians are crucial in overdose resuscitation and sustaining life of patients experiencing opioid overdose, prehospital providers may be uniquely responsible for understanding foundational systemic issues, such as racial inequity, and how they translate to emergency clinical care, which is not currently requisite by national EMS curriculum standards despite the measurable impact of care and access disparities experienced by patients of color **(95–105)**.

### Mental Health

It has been well established in the literature that the COVID-19 pandemic has correlated with worse mental health outcomes in healthcare workers (HCWs) **(20)**. The strongest correlation among COVID-19 and psychological symptoms appeared in female-identifying HCWs ages 30-49 years **(28)**. There is a continued need for strong psychological support and further investigation into determinants of poor mental health outcomes to develop evidence driven preventive measures.

### EMS Activation

911 dispatchers at public safety answering points (PSAPs) are critical to functional operations of the EMS system. Albright et. al investigated the efficacy of a dispatch screening tool to identify high suspicion of COVID-19 in patients based upon a questionnaire deployed by the 911 call taker in a study based in Massachusetts. The authors report a sensitivity of 74.9% (CI, 69.21-80.03) and a specificity of 67.7% (CI, 66.91-68.50) where n=263 **(47)**. Figure 2 is another example of a prehospital screening tool that may be helpful in supporting responding crew members in deciding necessary precautions and in clinical decision making. Amiry and Maguire report an exponential increase in emergency medical calls globally during the pandemic, so further investigation into ways PSAPs can best support responding clinicians using evidence based practices may be warranted **(21)**.

**Figure 2.**
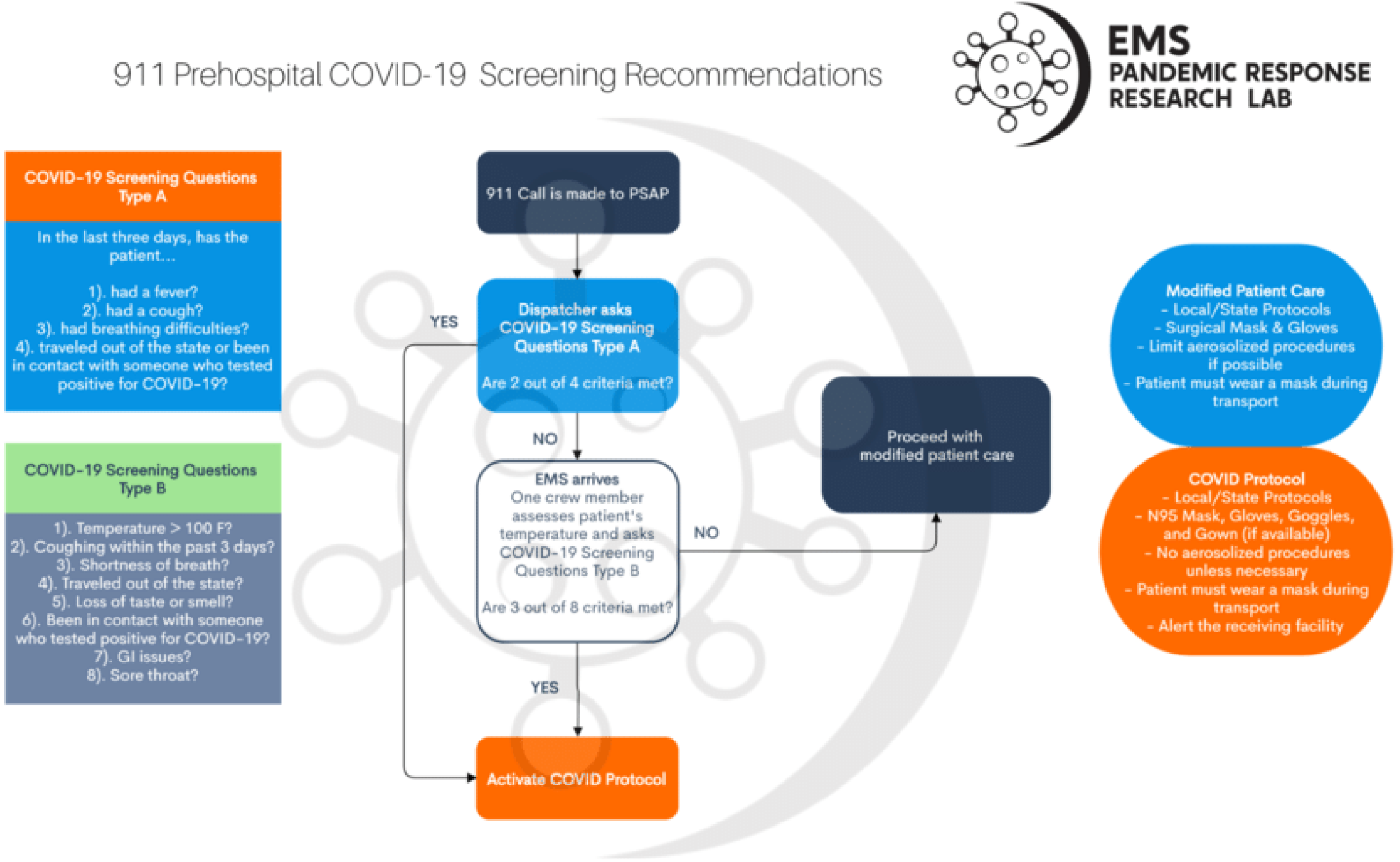
An example of standard 911 prehospital screening recommendations **(95)**. (*Permission for reproduction granted by the EMS Pandemic Response Research Lab*)

### Clinical Assessment, Treatment, and Outcomes

In a cohort study based in King County, Washington, US, Yang et. al found that EMS responded to approximately 16% of COVID-confirmed cases. From study initiation to June 2020 (four months), the study cohort mortality was 52.4%. Fever, tachypnea, and hypoxia was only present in a limited quantity of patients **(44)**. This is consistent with international prehospital consensus that suspicion of COVID-19 based solely on symptomatology should not be solely relied on for contemplating precautionary measures and isolation procedures. In a prehospital study in Denmark, patients under 60 years old were more likely to present with a loss of taste and/or smell upon assessment, however fever and cough were equally distributed across all age groups **(49)**. In England, Marincowitz et. al studied the accuracy of prognostic triage tools used prehospitally in the Yorkshire and Humber region. The WHO algorithm, PRIEST, NEWS2, and PMEWS tools all had high sensitivity for detecting COVID-19 infection **(65)**. In Uppsala, Sweden, Mälberg et. al report correlational findings of physiological respiratory parameters with patients suspected of having COVID-19. They found that the odds of COVID-19 diagnosis increased with respiratory rate, lower tidal volume, and negative inspiratory pressure **(65)**. In Paris, France, Jouffroy et. al report that pulse oximetry may be a reliable indicator for the need of mechanical ventilation in COVID-19 patients **(38)**. It is important to acknowledge, however, that clinically significant inaccuracy of pulse oximetry for detecting hypoxia in darker skin patients has been well established in the literature **(106,107)**. While many methods for assessing risk of COVID-19 infection exist, a universal guidance for prehospital assessment has not been established.

As COVID-19 is categorized as a respiratory illness, the majority of available articles on prehospital treatment had a significant focus on airway and breathing management. A study out of Paris reported that of the 15% of suspected COVID-19 patients who were found to be in cardiac arrest upon arrival, while the majority did not have prior cardiovascular risk factors present. This suggests that it is critical to provide sufficient respiratory support in the prehospital setting. It was found that of the suspected COVID-19 patients who were assessed by EMS personnel, 42% of BLS cases and 52% of ALS cases provided non-invasive ventilation, however, prehospital intubation was significantly less common as it was only present in 18% of ALS cases **(50)**. Jouffroy et al. report that of all COVID-19 suspected patients, 53.4% showed signs of respiratory distress. Most of these patients presented with respiratory rates of approximately 30/min and oxygen saturations of 72%. Additionally, all patients required some method of high flow oxygen treatment **(88)**

Studies, such as one conducted by Hart et al., provides recommendations for personal protection protocols for EMS providers. They suggest that if endotracheal intubation is needed, it should only be attempted once and with the assistance of video laryngoscopy to minimize time exposure to aerosols. A HEPA filter should then be added following the placement of an advanced airway to further minimize risk for contamination of the provider and all other equipment present in the ambulance. In the event that advanced airway intervention is not possible or clinically indicated, it is suggested to use two individuals for BVM ventilation to ensure that an air-tight seal is made with the mask. In agreement with other established protocols, they stress that donning of PPE should be performed before any contact is made with a suspected COVID-19 patient **(58)**.

As mentioned previously, there is a shortage of available resources not just pre-hospitality but in-hospital as well. Considerations need to be made to ensure resources are used in the most efficient manner. Natalzia et al. proposes a protocol to maximize cardiac arrest resources in EMS in which one should only initiate resuscitation of a patient in cardiac arrest if the arrest is witnessed or a shockable rhythm is present. They state that this protocol could account for 70.5% of favorable neurological outcomes and provide 6.3 additional available hospital beds per patient **(89)**.

### Education

As part of their initial education program, EMS students must generally acquire co-requisite field experience **(1)**. During the early era of the COVID-19 pandemic, it may have been difficult to participate in field experience opportunities due to facility policy limitations designed to reduce transmission. Grawey et. al examined an alternative model to the traditional ambulance “ride along” by providing a field-like experience in an emergency department setting. Grawey and colleagues report that this alternative experience was likely as effective as traditional prehospital clinical internships. Additionally, they found university medical students to have increased knowledge regarding EMS roles and responsibilities, which may have beneficial implications for transfer of care **(23)**. With regard to continuing education, the Commission on Accreditation for Prehospital Continuing Education (CAPCE) saw a 179% increase in EMS refresher courses during 2018 to 2020, with a 185% increase in asynchronous online learning **(54)**. This may suggest that distance learning as a method for recertifying provider credentials may increase rates of recertification, in contrast with in-person continuing education programs.

### Vaccine Perspectives and Prevalence

US EMS clinicians have also assumed nontraditional healthcare roles during the COVID-19 pandemic, such as in the case of a scope of practice expansion which allowed EMS clinicians to administer the COVID-19 to patients in states like New York, Massachusetts, and Vermont. Because EMS clinicians are highly trained in specific psychomotor clinical skills and knowledgeable in fundamental methods of patient assessment and treatment, it is likely that EMS in general has served unmeasurable utility through contributions in traditional and nontraditional roles. In April 2021, Gregory and colleagues conducted a cross sectional study of nationally certified EMS clinicians rostered with the National Registry of Emergency Medical Technicians (NREMT). Of a sample size of n=2584, 70% reported receiving a COVID-19 vaccine. Of the sampled population, 53% had concerns regarding the safety of COVID vaccines, 39% felt it was not necessary, and 84% of providers who did not receive any dose of the COVID vaccine did not plan to get it in the future **(25)**. The significant vaccine hesitancy amongst EMS clinicians poses a compelling public health concern, and approaches to mediate this, such as through local education and role modeling, may be necessary **(108)**.

### Limitations

Little literature exists on the interrogated area of investigation, and studies that are relevant to the area are often preliminary and may require continued study and design modifications to increase external validity. While all attempts to achieve a high level of internal validity were made as captured by robust quality assessments at each level of investigation, it was not possible to accommodate for every confounder. While this review is not exhaustive, the authors affirm satisfaction in the methods and results available at the time of study.

## CONCLUSION

US EMS clinicians are healthcare workers that have often held many roles and responsibilities during the COVID-19 pandemic. Within the subset domains of i. structural/systemic, ii. clinical outcomes, iii. clinical assessment, iv. treatment, v. special populations, vi. dispatch/activation, vii. education, viii. mental health, ix. perspectives/experiences, and x. transport, there is minimal high quality literature that faithfully reflect the status of modern American EMS in response to the COVID-19 pandemic. The available data reflect strides in contemporary approaches to assessment, treatment, transport, and education, but fall short in accounting for care and access disparities influenced by social determinants. Continued investigation on the impact of COVID-19 on EMS systems and personnel is warranted to ensure informed, appropriate, and evidence-based preparation for future pandemic and infectious disease response.

## Data Availability

All data produced in the present study are available upon reasonable request to the authors

## CONTRIBUTIONS

CAIV and EED gathered, extracted, and analyzed the data. CAIV, EED, JAD, LM, BJS, and RPL interpreted the data. CAIV, EED, and RBR wrote the first draft. JAD and EED assessed for methodical quality. BH was the PI of the lab, revised the first draft, and participated in revisions. All authors commented critically in the manuscript, revisions were made, and the final draft was prepared and submitted.

## INSTITUTIONAL DISCLAIMER

The work is solely that of the authors and does not necessarily represent the views, policies, or opinions of their affiliated institutions, employers, or partners. It was not reviewed or endorsed by any specific institution in particular.

## ACKNOWLEDGEMENTS

The authors would like to sincerely thank Paramedic Catherine Smith and EMT Rosa Turinetti for their expert opinions and experiences.

